# Influenza vaccination status ascertainment and vaccine effectiveness estimation: validity of self-report for current and prior season

**DOI:** 10.64898/2026.01.26.26344840

**Authors:** Rossana Peredo, Noémie Savard, Lea Separovic, Yuping Zhan, Rachid Amini, Marilou Kiely, Sara Carazo, Danuta M Skowronski

**Author notes:** **Corresponding author:** Noémie Savard, at : Direction des risques biologiques, Institut national de santé publique du Québec, 190 boul. Crémazie, Montréal, H2P 1E2, Canada.

## Abstract

Accurate vaccination status ascertainment is fundamental to valid vaccine effectiveness (VE) estimation. We evaluated the accuracy of current and prior season’s self-reported influenza vaccination status among outpatients with acute respiratory illness recruited by the Canadian Sentinel Practitioner Surveillance Network (SPSN) during the 2023-2024 season in Quebec, Canada. Vaccination status was self-reported at specimen collection for influenza virus testing and compared to the provincial vaccination registry. Self-report showed high registry agreement, including sensitivity, specificity, accuracy and kappa statistics for current (91%, 98%, 96%, 0.89) and prior (85%, 95%, 92%, 0.78) season’s vaccination status. Metrics were similar by influenza case status, age, sex and comorbidity with more variation for sensitivity than specificity. Test-negative design estimates of the crude association between influenza case status and current season’s self-reported or registry-based vaccination status did not meaningfully differ, with absolute difference of 2% overall. Findings support self-reported influenza vaccination for timely and valid VE estimation.

## Introduction

Timely and valid post-marketing vaccine effectiveness (VE) studies are needed to inform vaccination programs, especially those requiring seasonal vaccine reformulation, such as for influenza. As the core exposure variable, accurate vaccination status ascertainment is fundamental to valid VE estimation. In some settings, self-report within a few months of receipt may be the most reliable or only available source of vaccination information, notably when vaccine registries do not exist or are incomplete, unreliable or inaccessible. On that basis, the Canadian Sentinel Practitioner Surveillance Network (SPSN), the longest running platform for annual influenza VE estimation, has used self-reported influenza vaccination status consistently since its original pilot of the test-negative design (TND) methodology for this purpose in 2004 [1,2].

Several studies have examined the validity of influenza vaccine self-report [3–6], generally indicating high sensitivities (93-99%) but a wider range of specificities (65-97%) compared to health records or vaccine registries. The specificity of outcome ascertainment was previously shown to be more important than sensitivity for accurate influenza VE estimation, particularly in the context of lower ratios of influenza versus non-influenza virus attack rates [7]. Jackson *et al*. similarly showed through simulation that the specificity of vaccine status ascertainment is more important than sensitivity, with most pronounced misclassification impact on crude VE estimates in the context of low prevalence vaccine coverage [8]. Given potential influence on current season’s vaccine performance [9], some studies have also examined the validity of self-report for prior season’s influenza vaccination [5,10,11]. The aforementioned studies each pre-dated COVID-19, for which seasonal vaccination may now also further complicate accurate influenza vaccination recall.

The province of Quebec contributes to the Canadian SPSN and has an established influenza vaccination registry [12]. Using data collected from Quebec SPSN participants during the 2023-2024 season, we compared self-reported current and prior season’s influenza vaccination status with the provincial registry record, empirically exploring misclassification impact on crude TND estimates of the association between influenza vaccination and case status [2,13].

## Methods

### Setting and participants

The Canadian SPSN is a respiratory virus and VE monitoring network linking outpatient primary care providers in Canada’s four most populous provinces (Ontario, Quebec, British Columbia, Alberta). Patients with acute respiratory illness (ARI) presenting to SPSN sites are offered multiplex nucleic acid amplification testing for respiratory viruses (including influenza) by provincial public health laboratories. A questionnaire solicits influenza vaccine status and other information (e.g., sex, age, comorbidity) at specimen collection [1,14]. For this study, we included 2023-2024 SPSN participants ≥1 year, as per SPSN VE inclusion criteria, enrolled between October 29, 2023 and April 27, 2024 at any one of eight SPSN clinics in Quebec. Patients with distinct ARI episodes at least 14 days apart could contribute more than once.

### Vaccination status ascertainment

Consenting patients (or guardians) responded yes, no, or unknown to the questions: “Did patient receive the 2023-24 seasonal influenza vaccine” and “Did patient receive the 2022-2023 seasonal influenza vaccine (i.e. last season’s vaccine)?”. Self-report was compared to the provincial vaccination registry which includes all Québec residents and all vaccines administered in any setting. For the 2023-2024 season, registry-based vaccination was defined by record of receipt between September 1, 2023 and the corresponding ARI episode date. Prior season’s vaccination was based upon registry record between September 1, 2022 and February 28, 2023, the latest influenza vaccine registry record among Quebec SPSN participants.

### Statistical analysis

Agreement metrics compared self-report to registry-based influenza vaccination status including sensitivity, specificity, positive predictive value (PPV), negative predictive value (NPV), accuracy, and Cohen’s kappa coefficient, with 95% confidence intervals (CIs). Metrics were assessed for current and prior season’s vaccination, and further stratified by age group, self-reported comorbidity status (with/without) [15], and lab-reported influenza virus test result (test-positive case/test-negative control). Participants who reported ‘unknown’ current and/or prior season’s vaccination status were excluded from the missing season(s) analyses only.

Per usual TND approach, we compared the odds of being a test-positive case versus test-negative control among vaccinated versus non-vaccinated participants, summarizing via the odds ratio (OR) and deriving percent relative reductions among vaccinated individuals as (1-OR)x100. To assess the impact of vaccine status misclassification, we compared crude estimates based upon self-reported and registry-based vaccination information. Note that as per earlier simulations [8], but here using empirical data, we present crude estimates that cannot be interpreted as VE without incorporating additional considerations such as lag to vaccine effect and adjustment for confounding.

### Ethics

Conducted as a component of sentinel surveillance and program evaluation, this project obtained waiver of review from the Centre Hospitalier Universitaire de Québec-Université Laval research ethics board.

## Results

### Participant characteristics

Analyses included 2311 participants (ARI episodes), corresponding to 2279 distinct individuals, with median age 41 (interquartile range 19-59) years, 1437 (62%) female and 627 (27%) influenza test-positive (Table 1), similar to SPSN mid-season profile for all provinces [14]. Overall, <10% had unknown self-reported influenza vaccine status either season. We found all but two participants in the registry. Among those with known status, ∼20% self-reported current or prior season’s influenza vaccination, lowest among children 1-17 years (<10%) and increasing by age group, highest among adults >65 years (55-60%).

**Table 1.** Participant characteristics.

| Characteristics | All<br>N (%)<sup>a</sup> | Self-reported current season<br>influenza vaccination (2023-24); N (%)<sup>b</sup> |  |  | Self-reported prior season<br>influenza vaccination (2022-23); N (%)<sup>b</sup> |  |  |
| --- | --- | --- | --- | --- | --- | --- | --- |
|  |  | Yes,<br>(% among overall;<br>% among known) | No | Unknown | Yes,<br>(% among overall;<br>% among known) | No | Unknown |
| Total | 2311<sup>c</sup> (100) | 438 (19.0; 20.2) | 1732 (74.9) | 141 (6.1) | 493 (21.3; 23.4) | 1618 (70) | 200 (8.7) |
| **Age (years)** |  |  |  |  |  |  |  |
| Mean (SD) | 39.5 (24.0) | 56.9 (21.5) | 35.0 (22.5) | 40.2 (24.5) | 55.3 (22.3) | 34.4 (22.4) | 41.5 (24.1) |
| Median (IQR) | 41.0 (19-59) | 63.5 (44-72) | 37.0 (13-54) | 43 (17-59) | 62 (42-71) | 36 (13-53) | 46.0 (20-60) |
| **Age group** |  |  |  |  |  |  |  |
| 1-17 yr | 561 (24.3) | 34 (6.1; 6.5) | 490 (87.3) | 37 (6.6) | 46 (8.2; 8.9) | 468 (83.4) | 47 (8.4) |
| 18-49 yr | 849 (36.7) | 94 (11.1; 11.7) | 712 (83.9) | 43 (5.1) | 114 (13.4; 14.5) | 672 (79.2) | 63 (7.4) |
| 50-64 yr | 514 (22.2) | 110 (21.4; 23.0) | 368 (71.6) | 36 (7.0) | 122 (23.7; 26.5) | 338 (65.8) | 54 (10.5) |
| ≥ 65 yr | 387 (16.8) | 200 (51.7; 55.2) | 162 (41.9) | 25 (6.5) | 211 (54.5; 60.1) | 140 (36.2) | 36 (9.3) |
| **Influenza status** |  |  |  |  |  |  |  |
| Test-positive case |  |  |  |  |  |  |  |
| Overall | 627 (27.1) | 63 (10.0; 10.8) | 522 (83.3) | 42 (6.7) | 78 (12.4; 13.7) | 492 (78.5) | 57 (9.1) |
| 1-17 yr | 240 (38.3) | 11 (4.6; 4.9) | 215 (89.6) | 14 (5.8) | 16 (6.7; 7.3) | 204 (85) | 20 (8.3) |
| 18-49 yr | 227 (36.2) | 16 (7.0; 7.6) | 194 (85.5) | 17 (7.5) | 19 (8.4; 9.4) | 184 (81.1) | 24 (10.6) |
| 50-64 yr | 113 (18.0) | 20 (17.7; 19.0) | 85 (75.2) | 8 (7.1) | 22 (19.5; 21.4) | 81 (71.7) | 10 (8.8) |
| ≥ 65 yr | 47 (7.5) | 16 (34.0; 36.4) | 28 (59.6) | 3 (6.4) | 21 (44.7; 47.7) | 23 (48.9) | 3 (6.4) |
| Test-negative control |  |  |  |  |  |  |  |
| Overall | 1684 (72.9) | 375 (22.3; 23.7) | 1210 (71.9) | 99 (5.9) | 415 (24.6; 26.9) | 1126 (66.9) | 143 (8.5) |
| 1-17 yr | 321 (13.9) | 23 (7.2; 7.7) | 275 (85.7) | 23 (7.2) | 30 (9.3; 10.2) | 264 (82.2) | 27 (8.4) |
| 18-49 yr | 622 (26.9) | 78 (12.5; 13.1) | 518 (83.3) | 26 (4.2) | 95 (15.3; 16.3) | 488 (78.5) | 39 (6.3) |
| 50-64 yr | 401 (17.4) | 90 (22.4; 24.1) | 283 (70.6) | 28 (7.0) | 100 (24.9; 28.0) | 257 (64.1) | 44 (11.0) |
| ≥ 65 yr | 340 (14.7) | 184 (54.1; 57.9) | 134 (39.4) | 22 (6.5) | 190 (55.9; 61.9) | 117 (34.4) | 33 (9.7) |
| **Sex** |  |  |  |  |  |  |  |
| Female | 1437 (62.2) | 297 (20.7; 21.9) | 1058 (73.6) | 82 (5.7) | 332 (23.1; 25.2) | 988 (68.8) | 117 (8.1) |
| Male | 874 (37.8) | 141 (16.1; 17.3) | 674 (77.1) | 59 (6.8) | 161 (18.4; 20.4) | 630 (72.1) | 83 (9.5) |
| **Comorbidity** |  |  |  |  |  |  |  |
| With | 490 (21.2) | 195 (39.8; 41.0) | 281 (57.3) | 14 (2.9) | 205 (41.8; 43.9) | 262 (53.5) | 23 (4.7) |
| Without | 1755 (75.9) | 236 (13.5; 14.2) | 1429 (81.4) | 90 (5.1) | 282 (16.1; 17.4) | 1335 (76.1) | 138 (7.9) |
| Don't know | 66 (2.9) | 7 (10.6; 24.1) | 22 (33.3) | 37 (56.1) | 6 (9.1; 22.2) | 21 (31.8) | 39 (59.1) |

Abbreviations: SIQR: interquartile range; D, standard deviation; yr, years.
<sup>a</sup> Percentages are column-based, age values in years represent mean or median and parentheses represent SD or IQR, as specified
<sup>b</sup> Percentages are row-based, age values in years represent mean or median and parentheses represent SD or IQR, as specified.
<sup>c</sup> 2279 distinct individuals for 2311 episodes, including 30 individuals with two or more acute respiratory illness episodes.

### Agreement metrics

Tallies for self-reported relative to registry-based influenza vaccine status are shown in Table 2. Among those with known status, current season’s self-report had high overall accuracy (96%) including specificity (98%) and NPV (98%) with slightly lower sensitivity (91%) and PPV (90%) (Table 3). Metrics were only slightly lower for prior season, notably sensitivity (85%) and PPV (82%). Kappas showed strong agreement for current (0.89) and moderate for prior (0.78) season’s vaccination status [16].

**Table 2.**
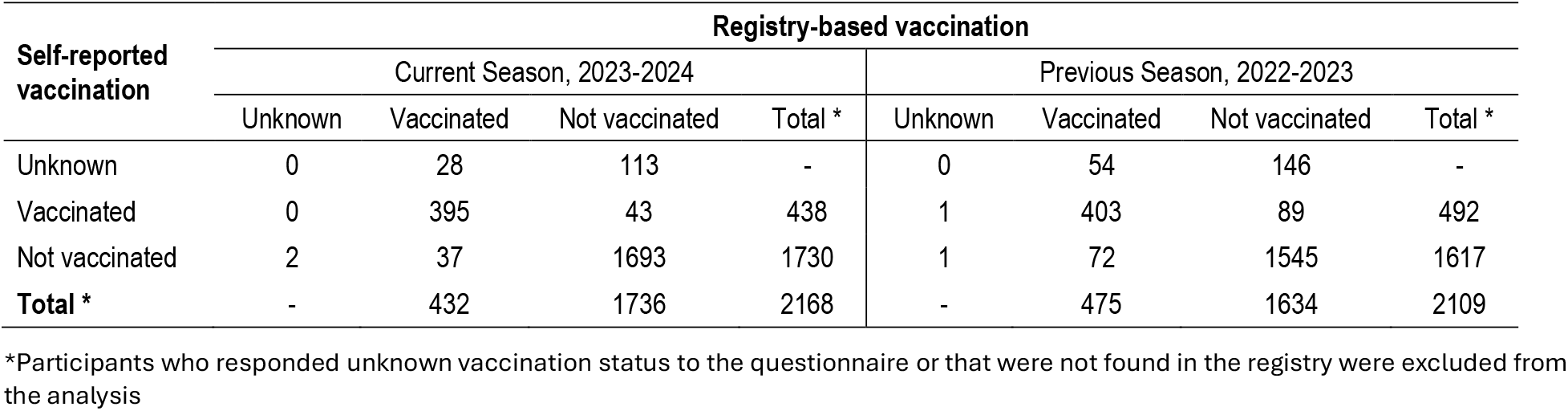
Current and prior season’s influenza vaccine status tallies, registry-based versus self-reported.

**Table 3.** Validity of self-reported versus registry-based influenza vaccination status, Quebec, 2023-24, current season vaccination.

| Characteristic | Current season (2023-2024) vaccine status metrics (95% CI) |  |  |  |  |  | Previous season (2022-2023) vaccine status metrics (95% CI) |  |  |  |  |  |
| --- | --- | --- | --- | --- | --- | --- | --- | --- | --- | --- | --- | --- |
|  | SN % | SP % | PPV % | NPV % | Acc % | Kappa | SN % | SP % | PPV % | NPV % | Acc % | Kappa |
| **Overall** | 91 (89-94) | 98 (97-98) | 90 (87-93) | 98 (97-99) | 96 (96-97) | 0.89 (0.86-0.91) | 85 (82-88) | 95 (93-96) | 82 (79-85) | 96 (95-97) | 92 (91-94) | 0.78 (0.75-0.82) |
| **Age group (years)** |  |  |  |  |  |  |  |  |  |  |  |  |
| 1-17 | 76 (62-91) | 98 (97-99) | 76 (62-91) | 98 (97-99) | 97 (95-98) | 0.75 (0.63-0.87) | 57 (43-72) | 96 (95-98) | 60 (46-74) | 96 (94-98) | 93 (91-95) | 0.55 (0.42-0.67) |
| 18-49 | 90 (83-96) | 98 (97-99) | 84 (77-91) | 99 (98-100) | 97 (96-98) | 0.85 (0.79-0.91) | 86 (79-93) | 95 (93-97) | 70 (62-79) | 98 (97-99) | 94 (92-96) | 0.74 (0.67-0.81) |
| 50-64 | 95 (91-99) | 97 (96-99) | 91 (86-96) | 99 (97-100) | 97 (95-98) | 0.91 (0.87-0.95) | 87 (81-93) | 94 (92-97) | 84 (77-90) | 96 (93-98) | 92 (90-95) | 0.80 (0.74-0.86) |
| ≥ 65 | 93 (89-96) | 94 (90-97) | 95 (92-98) | 91 (86-95) | 93 (91-96) | 0.86 (0.81-0.91) | 89 (85-93) | 87 (81-93) | 92 (88-96) | 83 (76-89) | 88 (85-92) | 0.75 (0.68-0.82) |
| **Influenza status** |  |  |  |  |  |  |  |  |  |  |  |  |
| Test-positive case |  |  |  |  |  |  |  |  |  |  |  |  |
| Overall | 85 (76-94) | 98 (96-99) | 81 (71-91) | 98 (97-99) | 96 (95-98) | 0.81 (0.73-0.89) | 85 (77-93) | 97 (96-99) | 82 (74-91) | 98 (97-99) | 96 (94-97) | 0.81 (0.74-0.88) |
| 1-17 yr | 78 (51-100) | 98 (96-100) | 64 (35-92) | 99 (98-100) | 97 (95-99) | 0.69 (0.45-0.92) | 75 (51-99) | 97 (94-99) | 56 (32-81) | 99 (97-100) | 95 (93-98) | 0.62 (0.40-0.84) |
| 18-49 yr | 73 (51-96) | 97 (95-100) | 69 (46-91) | 98 (96-99) | 96 (93-98) | 0.69 (0.49-0.88) | 89 (74-100) | 98 (97-100) | 84 (68-100) | 99 (97-100) | 98 (95-100) | 0.85 (0.72-0.98) |
| 50-64 yr | 87 (82-92) | 98 (95-100) | 90 (77-100) | 97 (96-98) | 96 (93-98) | 0.94 (0.85-1.00) | 84 (70-98) | 99 (96-100) | 95 (87-100) | 95 (90-100) | 95 (91-99) | 0.86 (0.75-0.98) |
| ≥65 yr | 83 (66-100) | 96 (88-100) | 94 (82-100) | 89 (77-100) | 91 (82-99) | 0.81 (0.63-0.99) | 90 (77-100) | 87 (73-100) | 87 (73-100) | 91 (79-100) | 88 (79-98) | 0.77 (0.58-0.96) |
| Test-negative control |  |  |  |  |  |  |  |  |  |  |  |  |
| Overall | 92 (89-95) | 97 (97-98) | 92 (89-95) | 98 (97-98) | 96 (95-97) | 0.89 (0.87-0.92) | 85 (81-88) | 93 (92-95) | 82 (78-85) | 95 (93-96) | 91 (90-93) | 0.77 (0.74-0.81) |
| 1-17 | 76 (59-93) | 99 (97-100) | 83 (67-98) | 98 (96-100) | 97 (95-100) | 0.77 (0.64-0.91) | 51 (35-68) | 96 (93-98) | 62 (44-80) | 94 (91-97) | 90 (87-94) | 0.51 (0.35-0.67) |
| 18-49 | 93 (87-99) | 98 (97-99) | 87 (80-95) | 99 (98-100) | 97 (96-99) | 0.89 (0.83-0.94) | 85 (77-93) | 94 (92-96) | 67 (58-77) | 98 (96-99) | 93 (91-95) | 0.71 (0.63-0.79) |
| 50-64 | 94 (89-99) | 97 (95-99) | 91 (85-97) | 98 (97-100) | 97 (95-98) | 0.90 (0.85-0.96) | 88 (81-95) | 93 (90-96) | 81 (73-89) | 96 (93-98) | 92 (89-94) | 0.79 (0.71-0.86) |
| ≥ 65 | 94 (90-97) | 93 (89-97) | 95 (92-98) | 91 (86-96) | 93 (91-96) | 0.86 (0.81-0.92) | 89 (85-93) | 87 (81-93) | 93 (89-96) | 81 (74-88) | 88 (85-92) | 0.75 (0.67-0.83) |
| **Sex** |  |  |  |  |  |  |  |  |  |  |  |  |
| Females | 91 (88-95) | 97 (96-98) | 90 (87-93) | 98 (97-98) | 96 (95-97) | 0.88 (0.85-0.91) | 87 (83-91) | 94 (93-96) | 83 (79-87) | 96 (95-97) | 93 (91-94) | 0.80 (0.76-0.84) |
| Males | 91 (87-96) | 98 (97-99) | 90 (85-95) | 98 (97-99) | 97 (96-98) | 0.89 (0.85-0.93) | 81 (75-87) | 95 (93-97) | 80 (74-86) | 95 (93-97) | 92 (90-94) | 0.75 (0.70-0.81) |
| **Comorbidity** |  |  |  |  |  |  |  |  |  |  |  |  |
| With | 96 (94-99) | 95 (93-98) | 93 (89-96) | 98 (96-99) | 96 (94-97) | 0.91 (0.87-0.95) | 91 (88-95) | 92 (89-94) | 89 (85-94) | 94 (91-96) | 92 (89-94) | 0.83 (0.78-0.88) |
| Without | 87 (83-91) | 98 (97-99) | 87 (83-92) | 98 (97-99) | 97 (96-97) | 0.86 (0.82-0.89) | 80 (76-85) | 95 (94-96) | 77 (72-82) | 96 (95-97) | 93 (91-94) | 0.74 (0.70-0.79) |

Abbreviations: Acc = accuracy; CI = confidence interval; NPV = negative predictive value; PPV = positive predictive value; SN = sensitivity; SP = specificity
Note: Metrics compare self-reported relative to registry-based vaccination status, the latter assumed to reflect true vaccination status. As per standard two-by-two table with registry-based columns and self-report-based rows; participants are considered vaccinated in cell “a” by both sources (true positives); in cell “b” by self-report but not registry (false positives); in cell “c” by registry but not self-report (false negatives); and in cell “d” by neither source (true negatives). Metrics derived as: $Accuracy = (a + d) / (a + b + c + d)$ ; $SN = a / (a + c)$ ; $SP = d / (b + d)$ ; $PPV = a / (a + b)$ ; $NPV = d / (c + d)$ . Levels of agreement by kappa interpreted as follows: ≤0.20 = none; 0.21–0.40 = minimal; 0.40–0.59 = weak, 0.60–0.79 = moderate, 0.80–0.90 = strong, and 0.91–1.00 = almost perfect agreement [16]

With age stratification, overall accuracy, specificity and NPV remained high among participants <65 years for both current (>97%) and prior (>92%) season, but all (notably NPV) were slightly lower among adults >65 years (>91% and >83%, respectively). Conversely, sensitivity and PPV metrics were lowest among 1-17-year-olds for both current (76% and 76%) and prior (57% and 60%) seasons, increasing by age group among adults for current (≥90% and ≥84%) and prior (>86% and >70%) seasons. Kappa estimates were also lowest among 1-17-year-olds for current (0.75) and prior (0.55) season. Accuracy, specificity and NPV did not meaningfully differ either season by influenza case status, sex or comorbidity with more variation observed for sensitivity and PPV. For current (but not prior) season, sensitivity and PPV were lower among influenza test-positive cases (85% and 81%) than controls (92% and 92%) including further stratified by age group (except 1-17-year-olds). Both sensitivity and PPV were lower among participants without (87% and 88%) than with (96% and 93%) comorbidity, more pronounced for prior season.

### Misclassification impact

Crude estimates of the association between current season’s influenza vaccination and case status were minimally biased with just 2% absolute difference between self-report and registry-based vaccination information (Figure 1). With stratification, CIs widen but absolute differences remain small, ranging 0-7% by age group, sex or comorbidity, except in the context of low vaccine coverage for 1-17-year-olds, with 16% absolute difference. Registry-based estimates tended higher, except in the context of the higher vaccine coverage among >65-year-olds, with self-report-based estimates instead 7% greater.

**Figure 1.**
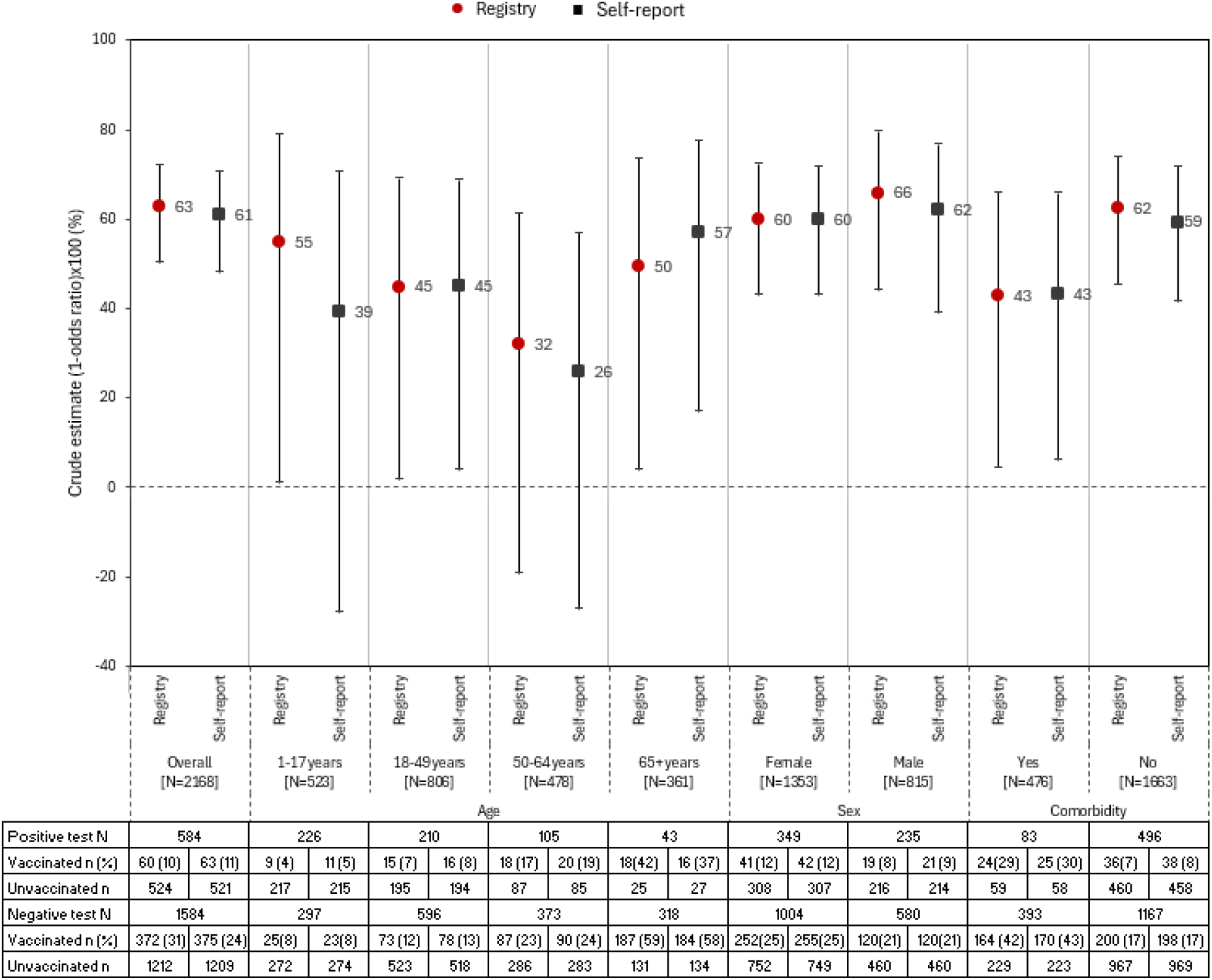
Crude association between influenza case and vaccination status, registry-based or self-report, Quebec, 2023-2024. Note: Crude estimates derived as (1-odds ratio) x 100% comparing the odds of being vaccinated versus the odds of being a case as per test-negative design but are only for illustration of the potential impact of misclassification of self-reported versus registry-based vaccine status, reflecting agreement metrics within the current data set. Estimates should not be interpreted as actual VE which requires consideration of other factors such as lag time (typically 14 days) for vaccine effect and adjustment for confounding, neither of which are incorporated here.

## Discussion

Among Quebec participants in the outpatient ARI surveillance and influenza VE monitoring platform of the Canadian SPSN, we report strong agreement between self-reported and registry-based ascertainment of influenza vaccination status, including 96% accuracy and 0.89 kappa for current season, slightly lower but still substantial for prior season at 92% accuracy and 0.78 kappa. While the impact of misclassification on VE estimates has been shown in previous simulation analyses to be influenced by specificity more than sensitivity, notably in the context of low vaccine coverage, we report both metrics to have been high for self-report versus registry record of current season’s influenza vaccination at 98% and 91%, respectively.

Our 2023-2024 metrics of self-report reliability are higher than other studies for specificity, ranging 66-97% elsewhere, and lower for sensitivity, ranging 92-99% elsewhere [3,5,6,17,18]. Context needs to be considered in comparing findings, including completeness of the reference record and noting ours is the first such evaluation post-COVID-19 pandemic. Registry-based misclassification is more likely to reflect false negative (i.e., failed documentation) than false positive vaccination record, potentially compromising the true specificity of self-report in relation to actual vaccine receipt. In that regard, our high specificity is consistent with high completeness of the Quebec registry where all vaccinators must report any vaccines given to any Quebec resident as a legal requirement, and with awareness-enhancing efforts during the pandemic. Conversely, our slightly lower sensitivity could reflect reduced or confused post-pandemic recall in the context of concomitant offer of influenza and COVID-19 vaccines.

A study by King *et al*. among 2014-2015 outpatients from a single US clinic also compared validity of self-report for prior versus current season’s influenza vaccination, showing virtually no difference for sensitivity (97% versus 98%) but lower prior season’s specificity (88% versus 97%); conversely, we report virtually no difference for specificity (95% versus 98%) but lower prior season’s sensitivity (85% versus 91%) [5]. In further stratified exploration, we found some variation by age group whereas King *et al*. found similar sensitivities for current season’s vaccination status, and others have reported high sensitivity (≥92%) even in children [5,17,18]. In the context of multiple childhood vaccine targets, some with similar sounding names (e.g., *Haemophilus influenzae*), and with sometimes simultaneous administration, poorer parental recall may not be unexpected, although likely to affect specificity more than sensitivity. A lower prevalence of influenza vaccine coverage in Quebec explains our lower PPV despite high specificity, and conversely high NPV despite lower sensitivity. Although publicly funded for any Quebec resident >6-months [19], in 2018 the Quebec Immunization Committee recommended targeted influenza vaccination for high-risk individuals only, with healthy >75-year-olds alone considered at heightened age-related risk [15]. Accordingly, the ∼6% of children and ∼20% of participants overall who self-reported current season’s 2023-2024 influenza vaccination in Quebec are ∼10% lower than overall SPSN report, even mid-season [14]. Other 2023-2024 surveys similarly show influenza vaccine coverage ∼10% lower in Quebec than the rest of Canada [20].

In combination with the likelihood of vaccination and influenza disease, validity metrics can be used to assess overall impact of vaccine status misclassification on VE estimates [8]. In general, non-differential misclassification is expected to bias toward a null association (i.e., OR closer to 1; 1-OR closer to zero), tending to underestimate VE. Our findings align with this expectation, indicating minimal misclassification impact, with self-report-based estimates of the crude association between vaccination and case status just 2% lower (absolute) than registry-based. Findings are similar with stratification by sex, comorbidity, and age among adults 18-64 years, with absolute under-estimation of 0-6% for self-report unlikely to meaningfully affect public policy decisions. We note two possible exceptions at the extremes of age that may also reflect instability with reduced sample size, including 17% (absolute) under-estimation by self-report among the least-vaccinated children 1-17 years; and conversely 7% (absolute) over-estimation by self-report among the most-vaccinated adults >65 years. The latter opposite direction of bias may reflect differentially lower case versus control sensitivity (83% versus 94%) in the context of lower NPV (89% versus 91%) compared to other age groups for whom NPV of self-reported vaccination status is >97% for both cases and controls.

Our study has limitations. Lack of vaccination registries limited analyses to one province but crude estimates of the association between influenza case and self-reported vaccination status in Quebec (61%) are similar to 2023-2024 SPSN estimates end-of-season overall (59%, not shown). However, findings may not be generalizable elsewhere and other factors beyond the scope of the current analysis can influence VE validity. Our results are limited to participants with known vaccination status, with <10% excluded on that basis. The provincial vaccination registry as comparator is not perfectly complete but given high specificity of self-report, missed recorded doses should be minimal. Finally, stratified analyses are limited by reduced sample size and wide CIs.

## Conclusion

Our findings reinforce the ongoing use of self-reported influenza vaccination status for timely and reliable seasonal influenza VE estimation by the longstanding Canadian SPSN and similar outpatient VE monitoring networks. Applicability to other seasonal vaccines (e.g., COVID-19) or settings warrants further evaluation.

## Data Availability

The institutional aggreements for data acces do not allow sharing individual participant data. Aggregated data in the present study are available upon reasonable request to the authors.

## Authorship contribution statement

**Rossana Peredo**: Writing – original draft, Writing – review & editing, Methodology, Formal analysis. **Noémie Savard**: Writing – original draft, Writing – review & editing, Methodology. **Lea Separovic**: Writing – review & editing, Methodology. **Yuping Zhan**: Writing – review & editing, Methodology. **Rachid Amini**: Writing – review & editing, Methodology. **Marilou Kiely**: Writing – review & editing, Methodology. **Sara Carazo**: Writing – review & editing, Methodology, Conceptualization. **Danuta M Skowronski**: Writing – review & editing, Methodology, Conceptualization.

## Acknowledgments

The authors wish to thank all the staff in the sentinel clinics, Josiane Rivard and Stéphanie Grenier from Centre de recherche du CHU de Québec-Université Laval (CR-CHUQ).

This project was supported by the ministère de la Santé et des Services sociaux du Québec and the Public Health Agency of Canada. The views expressed herein do not necessarily represent the views of these institutions. This work was also supported by the Fonds de recherche du Québec (FRQ) through the Research Centre grant.

